# Impact of Image Bit Depth Reduction on Deep Learning Performance in Chest Radiograph Analysis: A Multi-institutional Study

**DOI:** 10.64898/2026.03.07.26347853

**Authors:** Hirotaka Takita, Yasuhito Mitsuyama, Shannon L Walston, Kenichi Saito, Takahiro Sugibayashi, Masaki Okamoto, Chong Hyun Suh, Daiju Ueda

## Abstract

**Purpose:** Medical imaging typically generates 12- to 16-bit formats, yet conversion to 8-bit is often required. While deep learning has been widely explored in medical imaging, the influence of image bit depth on model performance is not fully understood. This study evaluates the impact of conversion from 16-bit to 8-bit for sex, age, and obesity classification using deep learning.

**Materials and methods:** In this retrospective, multi-institutional study, we analyzed 100,002 chest radiographs from 48,047 participants across three institutions. Three convolutional neural network architectures (ResNet52, EfficientNetB2, and ConvNeXtSmall) were trained on both 16-bit and 8-bit versions of the images. Model performance was evaluated using internal test datasets, randomly split multiple times, and an external test dataset. Statistical analysis included paired comparisons of area under the receiver operating characteristic curve (AUC-ROC) values, with Bonferroni correction for multiple comparisons.

**Results:** Across all architectures and classification tasks, differences between 16-bit and 8-bit model performance were minimal (mean differences ranging from -0.218% to 0.184%). Statistical analyses revealed no significant differences in AUC-ROC values between bit depths for any model-task combination (all p-values > 0.05 after Bonferroni correction). Effect sizes were small to moderate (Cohen’s d ranging from -0.415 to 0.391).

**Conclusion:** Reducing image bit depth from 16-bit to 8-bit does not significantly impact the performance of deep learning models in chest radiograph analysis. These findings suggest that 8-bit images can be used for deep learning applications in medical imaging without compromising model performance, potentially allowing for more efficient data storage and processing.

## Introduction

Medical imaging stands as an indispensable tool in modern healthcare, providing critical insights into patient anatomy and pathology that inform diagnosis, treatment planning, and disease monitoring. The integration of deep learning algorithms into medical image analysis has further revolutionized the field, enabling automated detection and classification tasks with unprecedented efficiency [1–4]. While various factors influence the performance of these algorithms, the role of image bit depth—the number of bits used to represent pixel intensity—has emerged as a subject of particular interest in the research community.

The conventional practice in medical imaging modalities such as radiography, computed tomography (CT), and magnetic resonance imaging (MRI) involves using higher bit depths (12-bit, 16-bit) to capture subtle tissue contrast variations. However, practical considerations often necessitate conversion to 8-bit formats, primarily due to storage efficiency requirements, computational resource optimization, and software compatibility constraints [5]. This transition between bit depths has historically raised theoretical concerns about potential information loss, particularly in the context of deep learning applications [6].

The implementation of deep learning models in medical imaging has prompted rigorous examination of various preprocessing parameters such as image resolution [7–9], and data augmentation strategies [10]. Yet comprehensive investigations into the specific implications of bit depth reduction have been relatively limited. This knowledge gap has motivated this systematic investigation into whether bit depth reduction meaningfully impacts model performance.

Our research methodology addresses this question through a comprehensive evaluation of deep learning model performance for chest radiograph analysis across different bit depth representations. By comparing the classification accuracy of convolutional neural networks using both 16-bit and 8-bit image formats, we investigate the impact of bit depth reduction on model performance. This study will provide insights into the practical implications of bit depth reduction in medical image analysis and potential optimization of future preprocessing protocols.

## Materials and methods Study Design

This retrospective, multi-institutional study was designed to evaluate the impact of image bit depth on deep learning model performance in medical imaging. Specifically, we compared the performance of models trained on 8-bit versus 16-bit PNG images for classifying three fundamental attributes: sex, age (elderly status), and obesity status. This approach allowed us to assess whether reducing bit depth from 16-bit to 8-bit affects model effectiveness. The study adhered to the principles outlined in the Declaration of Helsinki and received approval from the Ethics Committee of our University. The need for individual informed consent was waived, as the images were originally obtained during routine medical care and did not contain any patient-identifying information. We followed the STARD (Standards for Reporting of Diagnostic Accuracy) reporting guidelines to ensure transparency and comprehensiveness in our research methodology and findings [11].

## Source Dataset

The study used previously collected chest radiographs that included images and basic participant information such as sex, age, and body mass index (BMI) [12]. Data were collected from three collaborating institutions (Table 1). Institution A provided 66,796 images from 35,251 participants, comprising 15,877 males and 19,374 females, with an average age of 51 years and an average BMI of 23. Institution B contributed 18,899 images from 7,175 participants, including 3,720 males and 3,455 females, with an average age of 62 years and an average BMI of 23. Institution C supplied 14,307 images from 5,621 participants, consisting of 3,021 males and 2,600 females, with an average age of 59 years and an average BMI of 23.

**Table 1:** Demographics of the source dataset.

|  | Institution A | Institution B | Institution C |
| --- | --- | --- | --- |
| Splitting | Train/validation/internal test | Train/validation/internal test | External test |
| Images | 66796 | 18899 | 14307 |
| Participants | 35251 | 7175 | 5621 |
| Male | 15877 | 3720 | 3021 |
| Female | 19374 | 3455 | 2600 |
| Age (mean ± SD) | 51±11 | 62±13 | 59±13 |
| Age ≥ 65 | 9923 | 10110 | 6023 |
| Age < 65 | 56873 | 8789 | 8284 |
| BMI (mean ± SD) | 23±4 | 23±3 | 23±3 |
| BMI ≥ 25 | 15223 | 4154 | 3550 |
| BMI < 25 | 51573 | 14745 | 10757 |

BMI: body mass index

## Data Preparation and Image Processing

To conduct a rigorous analysis, data from Institutions A and B were combined to create an internal dataset for training the deep learning models, while data from Institution C served as an external test dataset to assess model generalizability [13]. For each image in the dataset, we prepared two versions: the original DICOM images were converted to 16-bit PNG format, and subsequently, an 8-bit PNG version was generated from the 16-bit files. To ensure robust evaluation of the models’ performance, we performed multiple random splits of the internal dataset into training, validation, and test sets (Figure 1). The number of splits was determined based on our power analysis, which indicated the required number of samples needed to detect meaningful differences between 8-bit and 16-bit model performance with sufficient statistical power. This approach allowed us to assess the consistency of our findings across different data distributions while maintaining the same underlying data quality.

**Figure 1:**
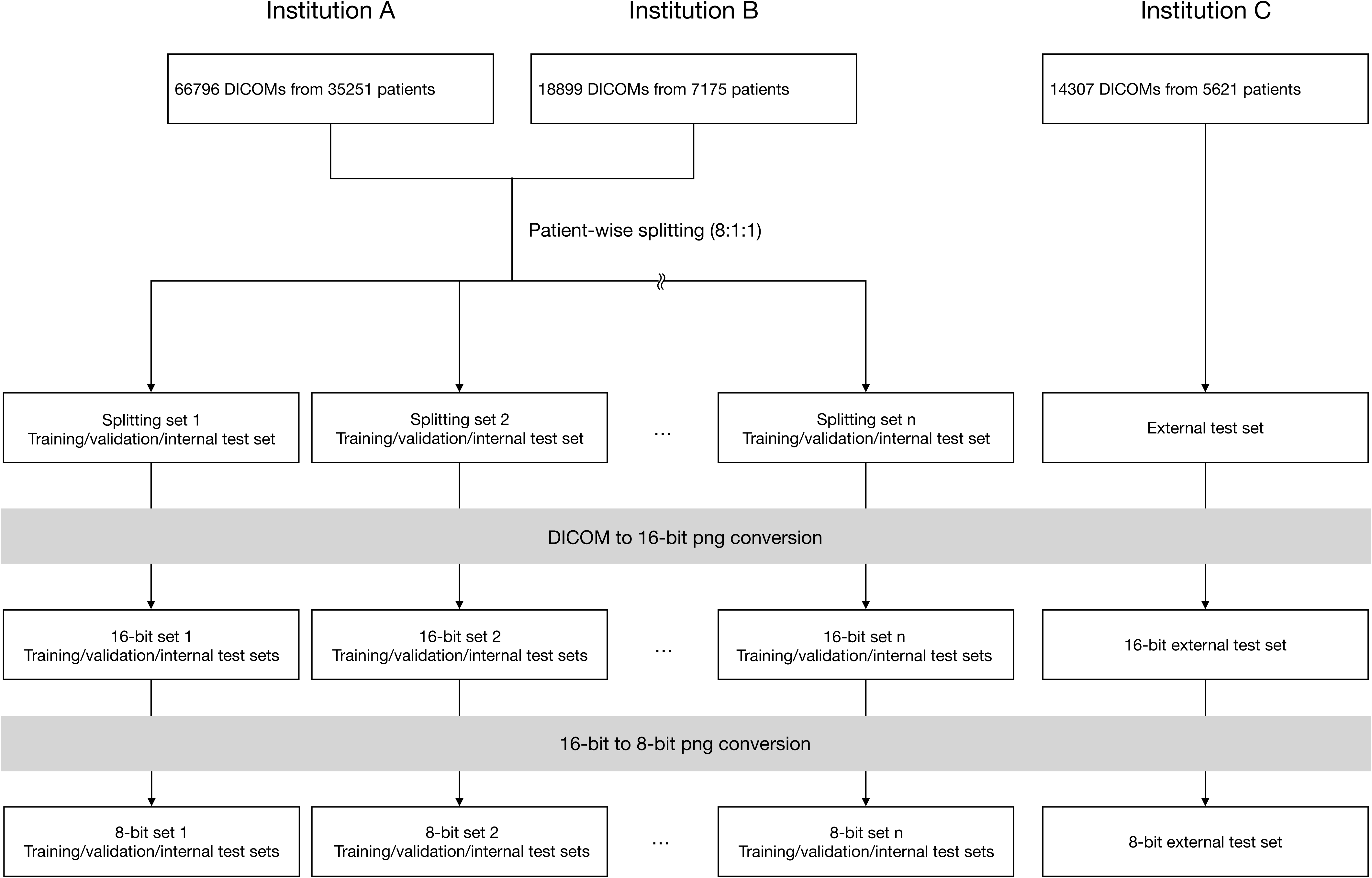
Flowchart of the data preparation and image processing. The flowchart illustrates the processing of DICOM images from three institutions (A: 66,796 DICOMs from 35,251 participants; B: 18,899 DICOMs from 7,175 participants; C: 14,307 DICOMs from 5,621 participants). Data from Institutions A and B were combined and underwent patient-wise splitting (8:1:1 ratio) into multiple training/validation/internal test sets, while Institution C data served as an external test set. All DICOM images were converted to 16-bit PNG format and subsequently to 8-bit PNG format, maintaining the split structure throughout the conversion process.

## Model Development

We developed AI models using three different architectures: ResNet52 [14], EfficientNetB2 [15], and ConvNeXtSmall [16]. Each architecture was trained separately on both 8-bit and 16-bit versions of the images to estimate sex, elderly status (age ≥65 years), and obesity (BMI ≥25). Cross-entropy loss was employed as the loss function. The models were trained from scratch using the training dataset and fine-tuned with the validation dataset, ensuring that all parameters were updated. The model iteration with the lowest loss value on the validation dataset, within a maximum of 100 epochs, was selected as the best-performing model.

For image preprocessing, the longest edge of each image was resized to 320 pixels while maintaining the original aspect ratio [8]. The shorter edge was padded with black pixels to reach 320 pixels, resulting in square images suitable for the input requirements of the deep learning architectures. No data augmentation techniques were applied. All development was carried out using PyTorch (version 2.0.1) [17]. A schematic of the model is provided in Online Resource 1, and the source code is available online [18].

## Model Testing

The classification performance of each model variant was assessed using both the internal test dataset (from Institutions A and B) and the external test dataset (from Institution C). Model performance was evaluated through sensitivity, specificity, accuracy, positive predictive value, negative predictive value, and area under the receiver operating characteristic curve (AUC-ROC). To quantify the uncertainty of the performance estimates, 95% confidence intervals for the AUC-ROC were calculated using the bootstrapping method. Each model was evaluated across all data splits to ensure robust performance assessment. The number of data splits used was determined by our power analysis to detect an AUC-ROC difference of 0.01 with 80% power at a significance level of 5%.

## Statistical Analysis

We hypothesized that there would be no significant difference in the performance of deep learning models trained on 16-bit images compared to those trained on 8-bit images for classifying sex, elderly status, and obesity from chest radiographs. Our aim was to statistically confirm the validity of using 8-bit images without compromising model performance.

Prior to the main analysis, we conducted a preliminary investigation to estimate the variability of the AUC-ROC differences between models trained on 16-bit and 8-bit images. This investigation revealed that the standard deviation (SD) of the AUC-ROC differences was at most 0.01 in all trials. So, we assumed an effect size (mean difference in AUC-ROC) of 0.01 as the smallest meaningful difference. The power analysis indicated that a sample size of 10 pairs (i.e., 10 data splits) would be sufficient to detect differences with 80% power at the 0.05 significance level.

For the statistical comparison between the 16-bit and 8-bit models, we conducted paired analyses of the AUC-ROC for each model-task combination. The differences in AUC-ROCs (calculated as 16-bit minus 8-bit) across the 10 data splits were used to assess the effect of bit depth. To evaluate the normality of the distribution of these AUC-ROC differences, we employed the Shapiro-Wilk test. If the AUC-ROC differences were found to be normally distributed (p > 0.05), we used a paired t-test to compare the means. If normality was not met (p ≤ 0.05), we employed the non-parametric Wilcoxon signed-rank test. Given that multiple comparisons were made across the three models and three tasks—a total of nine tests—we applied the Bonferroni correction to control for the family-wise error rate. Effect sizes were calculated using Cohen’s d for paired samples to quantify the magnitude of any differences between the 16-bit and 8-bit models. All statistical analyses were conducted using Python 3.9 and the SciPy library (version 1.5.2). The stats module was used for performing the Shapiro-Wilk test, paired t-tests, and Wilcoxon signed-rank tests.

## Results

Our evaluation compared three deep learning models (ResNet, EfficientNet, and ConvNeXt) across three classification tasks (Sex, Elderly, and Obesity) using both 8-bit and 16-bit settings. The models were tested on data from three institutions (A, B, and C), with results averaged over 10 runs (Table 2). For Sex classification, all models achieved perfect performance (AUC-ROC 1.00) regardless of bit depth or institution. In the Elderly classification task, performance was strong but showed some variation across institutions: Institution A achieved AUC-ROCs of 0.95-0.96, Institution B showed slightly lower performance with AUC-ROCs of 0.94-0.95, and Institution C demonstrated the lowest performance with AUC-ROCs of 0.91-0.93. EfficientNet marginally outperformed other models in this task, achieving the highest AUC-ROC of 0.96 at Institution A. For Obesity classification, all models showed robust performance, with Institution A achieving the highest AUC-ROCs (0.97-0.98), followed by Institution B (0.96-0.97), and Institution C (0.94-0.95). Notably, there were no substantial differences in performance between 8-bit and 16-bit across any of the tasks or models, suggesting that 8-bit could be implemented without significant performance degradation.

**Table 2:** The results of area under the receiver operating characteristic curves in each institution.

| Task | Model | Bit depth | Institution A | Institution B | Institution C |
| --- | --- | --- | --- | --- | --- |
|  |  |  | Internal test | Internal test | External test |
| Sex | ResNet | 8bit | 1.00(1.00-1.00) | 1.00(1.00-1.00) | 1.00(1.00-1.00) |
|  |  | 16bit | 1.00(1.00-1.00) | 1.00(1.00-1.00) | 1.00(1.00-1.00) |
|  | EfficientNet | 8bit | 1.00(1.00-1.00) | 1.00(1.00-1.00) | 1.00(1.00-1.00) |
|  |  | 16bit | 1.00(1.00-1.00) | 1.00(1.00-1.00) | 1.00(1.00-1.00) |
|  | ConvNeXt | 8bit | 1.00(1.00-1.00) | 1.00(1.00-1.00) | 1.00(1.00-1.00) |
|  |  | 16bit | 1.00(1.00-1.00) | 1.00(1.00-1.00) | 1.00(1.00-1.00) |
| Elderly | ResNet | 8bit | 0.95(0.95-0.96) | 0.94(0.93-0.95) | 0.93(0.92-0.93) |
|  |  | 16bit | 0.95(0.95-0.96) | 0.95(0.94-0.96) | 0.93(0.92-0.93) |
|  | EfficientNet | 8bit | 0.96(0.95-0.96) | 0.95(0.94-0.96) | 0.93(0.93-0.94) |
|  |  | 16bit | 0.96(0.95-0.96) | 0.95(0.94-0.96) | 0.93(0.93-0.94) |
|  | ConvNeXt | 8bit | 0.95(0.94-0.95) | 0.94(0.93-0.95) | 0.92(0.91-0.92) |
|  |  | 16bit | 0.95(0.94-0.96) | 0.94(0.92-0.95) | 0.91(0.91-0.92) |
| Obesity | ResNet | 8bit | 0.98(0.97-0.98) | 0.97(0.96-0.97) | 0.94(0.93-0.94) |
|  |  | 16bit | 0.98(0.97-0.98) | 0.97(0.96-0.97) | 0.94(0.94-0.94) |
|  | EfficientNet | 8bit | 0.98(0.97-0.98) | 0.97(0.96-0.97) | 0.94(0.94-0.95) |
|  |  | 16bit | 0.98(0.97-0.98) | 0.97(0.96-0.98) | 0.95(0.94-0.95) |
|  | ConvNeXt | 8bit | 0.98(0.97-0.98) | 0.96(0.95-0.97) | 0.95(0.94-0.95) |
|  |  | 16bit | 0.97(0.97-0.98) | 0.96(0.95-0.97) | 0.95(0.94-0.95) |

Our comparison of deep learning models trained on 16-bit versus 8-bit chest radiographs revealed minimal differences across all classification tasks and architectures (Figure 2). For sex classification, all models showed negligible mean differences (ResNet: 0.007% ± 0.042%, EfficientNet: -0.002% ± 0.014%, ConvNeXt: 0.010% ± 0.131%). Elderly status classification demonstrated slightly larger but still insignificant differences (ResNet: -0.103% ± 0.732%, EfficientNet: 0.076% ± 0.542%, ConvNeXt: -0.218% ± 0.525%). Similarly, obesity classification maintained minimal differences (ResNet: 0.160% ± 0.443%, EfficientNet: 0.184% ± 0.470%, ConvNeXt: -0.006% ± 0.547%).

**Figure 2:**
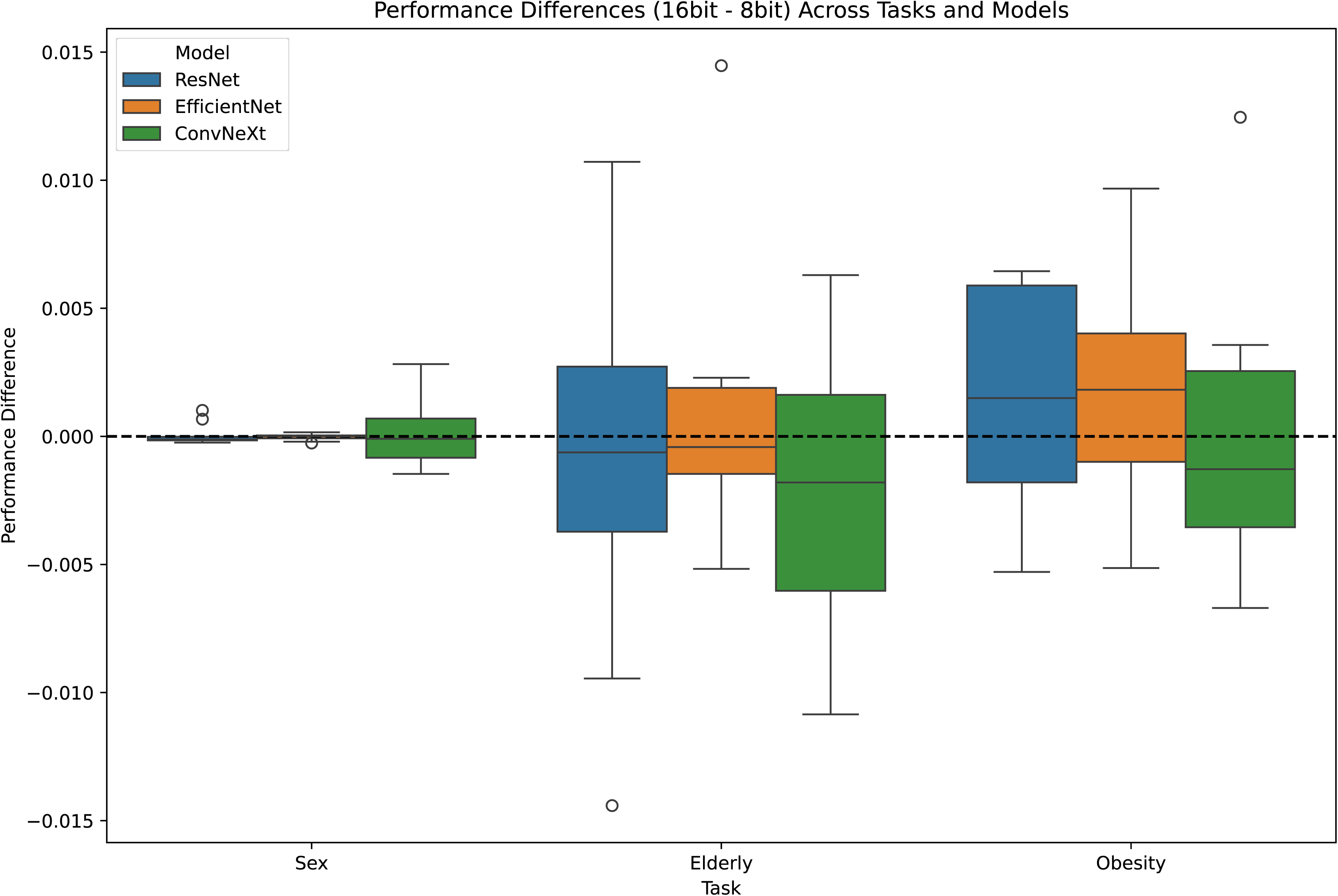
Performance differences between 16-bit and 8-bit image-based deep learning models across classification tasks. Box plots show the performance differences (16-bit minus 8-bit) for three deep learning architectures (ResNet, EfficientNet, and ConvNeXt) across three classification tasks (sex, elderly status, and obesity) using chest radiographs. The horizontal dashed line at y=0 represents no difference between bit depths. Boxes indicate interquartile ranges, whiskers extend to 1.5 times the interquartile range, and circles represent outliers. All models demonstrated minimal performance differences (<0.2%) between 16-bit and 8-bit images across all tasks, suggesting bit depth reduction had negligible impact on model performance.

Statistical analyses were conducted to compare the performance of models trained on 16-bit versus 8-bit images across three classification tasks (sex, elderly status, and obesity) and three deep learning architectures (ResNet, EfficientNet, and ConvNeXt) (Table 3). For sex classification, the Shapiro-Wilk test indicated non-normal distribution for ResNet (p = 0.0005), while EfficientNet and ConvNeXt showed normal distributions (p = 0.345 and p = 0.483, respectively). The Wilcoxon signed-rank test for ResNet (W = 19.0, p = 0.432) and paired t-tests for EfficientNet (t = -0.423, p = 0.682) and ConvNeXt (t = 0.234, p = 0.820) revealed no significant differences between 16-bit and 8-bit models. Effect sizes were small for all models (Cohen’s d: ResNet = 0.165, EfficientNet = -0.134, ConvNeXt = 0.074). For elderly status classification, the Shapiro-Wilk test indicated non-normal distribution for EfficientNet (p = 0.014), while ResNet and ConvNeXt showed normal distributions (p = 0.881 and p = 0.971, respectively). The paired t-test for ResNet (t = -0.445, p = 0.667) and ConvNeXt (t = -1.314, p = 0.221), and the Wilcoxon signed-rank test for EfficientNet (W = 27.0, p = 1.000) showed no significant differences between bit depth models. Effect sizes were small to moderate (Cohen’s d: ResNet = -0.141, EfficientNet = 0.140, ConvNeXt = -0.415). For obesity classification, all models showed normal distributions (ResNet: p = 0.163, EfficientNet: p = 0.922, ConvNeXt: p = 0.172). Paired t-tests revealed no significant differences between 16-bit and 8-bit models for ResNet (t = 1.139, p = 0.284), EfficientNet (t = 1.237, p = 0.247), or ConvNeXt (t = -0.035, p = 0.973). Effect sizes ranged from negligible to moderate (Cohen’s d: ResNet = 0.360, EfficientNet = 0.391, ConvNeXt = -0.011).

**Table 3:** The results of the statistical analysis in the external test dataset.

| Task | Model | AUC-ROCs |  | Test | Statistic | p-value |
| --- | --- | --- | --- | --- | --- | --- |
|  |  | AUC-ROCs<br>difference (mean) | AUC-ROCs<br>difference<br>(median) |  |  |  |
| Sex | ResNet | 0.00007 | -0.00012 | Wilcoxon<br>signed-rank test | 19.00 | 0.43 |
|  | EfficientNet | -0.00002 | -0.00062 | Paired t-test | -0.42 | 0.68 |
|  | ConvNeXt | 0.00010 | 0.00150 | Paired t-test | 0.23 | 0.82 |
| Elderly | ResNet | -0.00103 | 0.00001 | Paired t-test | -0.45 | 0.67 |
|  | EfficientNet | 0.00076 | -0.00042 | Wilcoxon<br>signed-rank test | 27.00 | 1.00 |
|  | ConvNeXt | -0.00218 | 0.00182 | Paired t-test | -1.31 | 0.22 |
| Obesity | ResNet | 0.00160 | -0.00009 | Paired t-test | 1.14 | 0.28 |
|  | EfficientNet | 0.00184 | -0.00180 | Paired t-test | 1.24 | 0.25 |
|  | ConvNeXt | -0.00006 | -0.00128 | Paired t-test | -0.04 | 0.97 |
| AUC-ROCs: area under the receiver operating characteristic curves |  |  |  |  |  |  |

## Discussion

The primary objective of this study was to evaluate the impact of image bit depth on the performance of deep learning models in medical imaging, specifically in the classification of sex, elderly status, and obesity from chest radiographs. Our findings indicate that reducing the bit depth from 16-bit to 8-bit does not significantly affect the performance of deep learning models across multiple architectures. Statistical analyses for each model-task combination revealed negligible differences in the AUC-ROCs, with all p-values exceeding the threshold. These results suggest that the theoretical concerns regarding information loss during bit depth reduction may not have practical implications for deep learning applications in medical imaging. The minimal differences observed across models and tasks support the notion that 8-bit images are sufficient for training effective deep learning models in this context.

While prior research has investigated the influence of various image preprocessing techniques—such as resolution adjustments and data augmentation—on deep learning model performance [19–21], the specific effect of bit depth has not been thoroughly examined in the context of classification tasks on radiological images.

However, a related study on nuclei instance segmentation in fluorescence-stained histological images found that models trained on 8-bit images performed comparably to those trained on 16-bit images [22]. Our study extends these findings to classification tasks in chest radiography, this suggests that the critical features necessary for deep learning models may be preserved despite bit depth reduction across different medical imaging tasks and modalities.

The practical implications of our findings are substantial for the field of medical imaging [23]. The use of 8-bit images can lead to significant reductions in storage requirements and computational resources, facilitating more efficient data management and model training processes [24]. This is particularly relevant for institutions with limited storage capacity or computational power. Moreover, the compatibility of 8-bit images with a wider range of software tools and platforms can enhance collaboration and data sharing among researchers and clinicians. By demonstrating that 8-bit images do not compromise model performance, our study supports the adoption of more streamlined preprocessing protocols without sacrificing diagnostic accuracy.

Despite the robust methodology and analysis, our study has several limitations. First, the classification tasks were limited to sex, elderly status, and obesity, which may not capture the complexity of other diagnostic challenges in medical imaging, such as detecting subtle pathological changes. Future studies should explore the impact of bit depth on models tasked with identifying specific diseases or abnormalities. Second, the dataset comprised chest radiographs from three institutions, which may not represent all variations encountered in clinical practice [25]. Third, we did not incorporate other modalities such as CT scans, MRI, or ultrasound images, which might exhibit different sensitivity to bit depth reduction given their distinct characteristics and information content.

Building upon our findings, future research should investigate the effect of bit depth reduction on more complex tasks. Considering the positive results from both our study and the related work on nuclei instance segmentation, there is potential for broader application of 8-bit images in different medical imaging contexts.

Additionally, exploring other imaging modalities could provide a more comprehensive understanding of how bit depth influences deep learning models. Evaluating the interplay between bit depth and model interpretability or explainability could further enhance the clinical applicability of deep learning models.

Our study demonstrates that reducing image bit depth from 16-bit to 8-bit does not significantly affect the performance of deep learning models in classifying sex, elderly status, and obesity from chest radiographs. These findings support the use of 8-bit images in medical imaging deep learning applications, offering potential benefits in storage efficiency, computational resource optimization, and software compatibility without compromising diagnostic accuracy. The results encourage a reevaluation of standard preprocessing protocols, promoting more efficient and accessible deployment of deep learning models in clinical settings.

## Data Availability

All data produced in the present study are available upon reasonable request to the authors

## Acknowledgements

We used ChatGPT (GPT-5.4 Pro, accessed 7 March 2026, OpenAI, San Francisco, CA, USA) for assistance with parts of the English proofing.

## Ethical statement

The study adhered to the principles outlined in the Declaration of Helsinki and received approval from the Ethics Committee of Osaka Metropolitan University. The need for individual informed consent was waived, as the images were originally obtained during routine medical care and did not contain any patient-identifying information.

## Data sharing statement

The data used and analyzed during the current study are available from the corresponding author on reasonable request.

## Authors’ contribution statements

Daiju Ueda contributed to the study conception and design. Material preparation and data collection were performed by Yasuhito Mitsuyama and Daiju Ueda. Analysis was performed by Daiju Ueda. The draft of the manuscript was written by Hirotaka Takita and Daiju Ueda, and all authors commented on previous versions of the manuscript. All authors read and approved the final manuscript.

