## supplemental file for "Impact of Image Bit Depth Reduction on Deep Learning Performance in Chest Radiograph Analysis: A Multi-institutional Study"

**Table of Contents:**

**Section S1: Supplemental Figures**

Supplemental Figure 1: Model overview

**Section S2: References for the Supplementary Appendix**

**Section S1: Supplemental Figures**
**Supplemental Figure 1: Model overview**

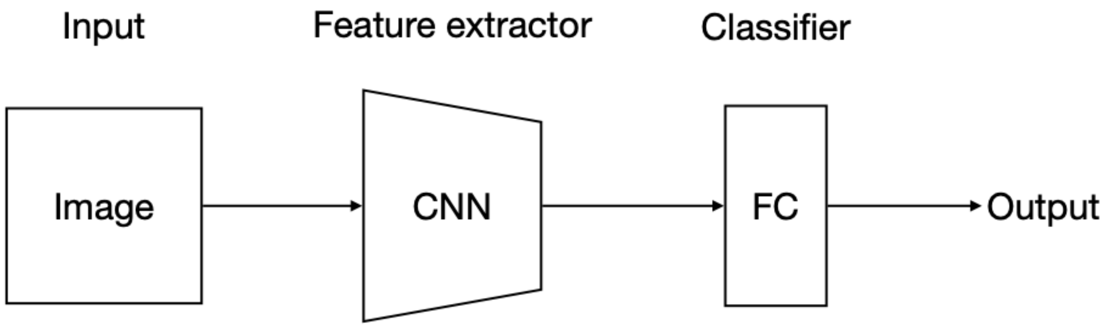


The deep learning models used in this study comprised convolutional neural networks (CNNs) - specifically ResNet52 [1], EfficientNetB2 [2], and ConvNeXtSmall [3] - as feature extractors and fully connected layers (FC) as classifiers. The chest radiographs are fed to each CNN block, where they are downsampled each time they are passed through the convolution layer. The results of these CNN blocks are then passed to their corresponding classifiers.
